# Mathematical Analysis of COVID-19 Transmission Dynamics with a Case Study of Nigeria and its Computer Simulation

**DOI:** 10.1101/2020.10.20.20216473

**Authors:** B. C. Agbata, Ogala Emmanuel, Tenuche Bashir, Obeng-Denteh William

## Abstract

In this article, we formulated a mathematical model for the spread of the COVID-19 disease and we introduced quarantined and isolated compartments. The next generation matrix method was adopted to compute the basic reproduction number (*R*_0_) in order to assess the transmission dynamics of the COVID-19 deadly disease. Stability analysis of the disease free equilibrium is investigated based on the basic reproduction number and the result shows that it is locally and asymptotically stable for *R*_0_ less than 1. Numerical calculation of the basic reproduction number revealed that *R*_0_ < 1 which means that the disease can be eradicated from Nigeria. The study shows that isolation, quarantine and other government policies like social distancing and lockdown are the best approaches to control the pernicious nature of COVID-19 pandemic.

## 1. Introduction

Mathematical model and its computer simulations are great tools that can be used to control and predict infectious diseases globally. The Covid-19 pandemic which is popularly known as coronavirus pandemic, a recent pandemic caused by severe acute respiratory syndrome coronavirus (SAR-Covid-2). The new virus was named by International committee on Taxonomy of viruses (ICTV) as “SARS-Covid-2” because it is generally related to the coronavirus responsible for the SARS outbreak of 2003 [4,5]. The recent pandemic was first identified in December 2019 in Wuhan China and it was declared to be a Public Health Emergency of International concern.

On 30 January, 2020 and as pandemic on 11 March 2020 [4]. As of 19 September 2020, the disease was confirmed in more than 30.3 million cases reported globally in 188 countries and territories with over 949,000 deaths and over 206 million have fully recovered globally [10].

The pandemic was first confirmed and announced in Nigeria on 27 February, 2020, when an Italian citizen in Lagos tested positive for the virus and the second case was reported on 9 March, 2020 in Ewekoro, Ogun State when a Nigerian citizen had contact with the Italian citizen [1,3,6].

Recently, the spread of covid-19 in Nigeria increases continually as the latest record provided by the Nigeria Centre for Disease Control declares Nigeria to reveal a total of 56,956 confirmed cases [2]. On 18 September 2020, Nigeria recorded 221 new confirmed cases and 1 death out of a total of daily test of 2,609 samples carried out across the country, the 221 new cases reported from 18 state – Lagos (59), Abia (46), FCT (22), Gombe (20), Plateau (17), Rivers (11), Bauchi (7), Benue (6), Ekiti (6) Imo (6), Kaduna (4), Kwara (4), Ondo (4), Ogun (3), Osun (3), Bayelsa (1), Edo (1), Kano (1) and these results bring Lagos state total confirmed cases to 18,827, followed by Abuja (5,526), Oyo (3,226), Plateau (3,192) Edo (2,611), Kaduna (2,326), Rivers (2,220), Delta (1,799), Ogun (1,758), Kano (1,734), Ondo (1,594), Enugu (1,234), Ebonyi (1,035), Kwara (1,013), Abia (881), Kastina (845), Osu (810), Gombe (799), Borno (741), Bauchi (689), Imo (557), Benue (473), Nasarawa (447), Bayelsa (394), Jigawa (322), Ekiti, (313), Akwa Ibom (228), Niger (258), Anambra (232), Adamawa (230), Sokoto (161), Taraba (95), Yobe (73), and Kogi (3) [7].

The Covid-19 outbreak is seen as the greatest global threat worldwide because it spreads globally and millions of confirmed cases, accompanied by thousands deaths globally. As of 14 September 2020, WHO reports 28,637,952 confirmed cumulative cases with 917,417 deaths in the world and 18 September 2020, total of 56,956 confirmed cases, total of 48,305 discharged and total deaths of 1,094 occurred in Nigeria [9].

Covid-19 is spread primarily from human to human through direct or indirect close proximity with contaminated surface, objects or with secretion droplets, saliva or respiratory secretions expelled from mouth or nose of an infected individual coughs, sings, sneezes or speaks.

The sign and symptoms of Covid-19 usually start two to 14 days after contact with infected person or contaminated surface, this period of time after exposure and before having symptoms is known as incubation period. Common sign and symptoms of Covid-19 include, cough, fever, tiredness, shortness of breath, sore throat etc. Older people with underlying medical challenges like serious heart diseases (heart failure, coronary arterial disease or cardionryopatty), cancer, type 2 diabetes, sickle cell disease, high blood pressure[14].

Mathematical modelling and its computer simulation has attracted the attention of many researchers because of its striking applications in the area of prediction and control of infectious diseases. Many researchers have proposed mathematical models for prediction of infectious disease. [15], Proposed Mathematical Assessment of the transmission dynamics of HIV/AIDs with treatment effects. Their model comprises of four compartments considering treatment class as the control measure. [12], proposed a mathematical model for simulating the phase-based transmissibility of a novel Covid-19.

In their study, they developed a Bats-Hosts Reservoir-people transmission network model for simulating the potential transmission from source of infection to the human infection. The basic reproduction number from reserviour to person analyzed was estimated to be 2.30 and 3.58 from person to person.

Their model revealed that the transmissibility of SARS-COV-2 was higher than the middle east respiratory syndrome within the Middle East countries. Our main contribution in this article is addition of new compartments such as quarantine, isolation and death which are not considered in most existing models, the addition of these compartments give rise to a number of analysis that can be used to study COVID-19 transmissibility intensively.

## 2.0 Mathematical Formulation Of The Model

In this section, we developed a mathematical model on COVID-19 based on some realistic mathematical assumptions about Covid-19. The total population is subdivided into seven compartments, namely susceptible *S*(*t*), exposed E(t), quarantine Q(t), infected (*I*_*n*_), isolate *I*_*s*_(*t*), Death *D*(*t*) and recovered *R*(*t*) let *λ* be the constant recruitment rate to the susceptible population, the rate at which people become susceptible to the disease and m the rate which the susceptible individuals become exposed to the disease, *β* is the rate of infection. *ω* and *α* are the rate at which the quarantine individuals because susceptible and susceptible individuals are removed from the population respectively. The exposed individuals move to the quarantine class through the rate b and get recovered through the rate *a*.

The isolated individuals get recovered at the rate *k* and *μ*_2_ is disease induced death rate. Quarantined individuals get infected at the rate *θ* and infected individual recovered through the rate *d*, the natural death is *μ* in every compartment. Figure 1 gives the schematic diagram of the model.

**Figure 1.**
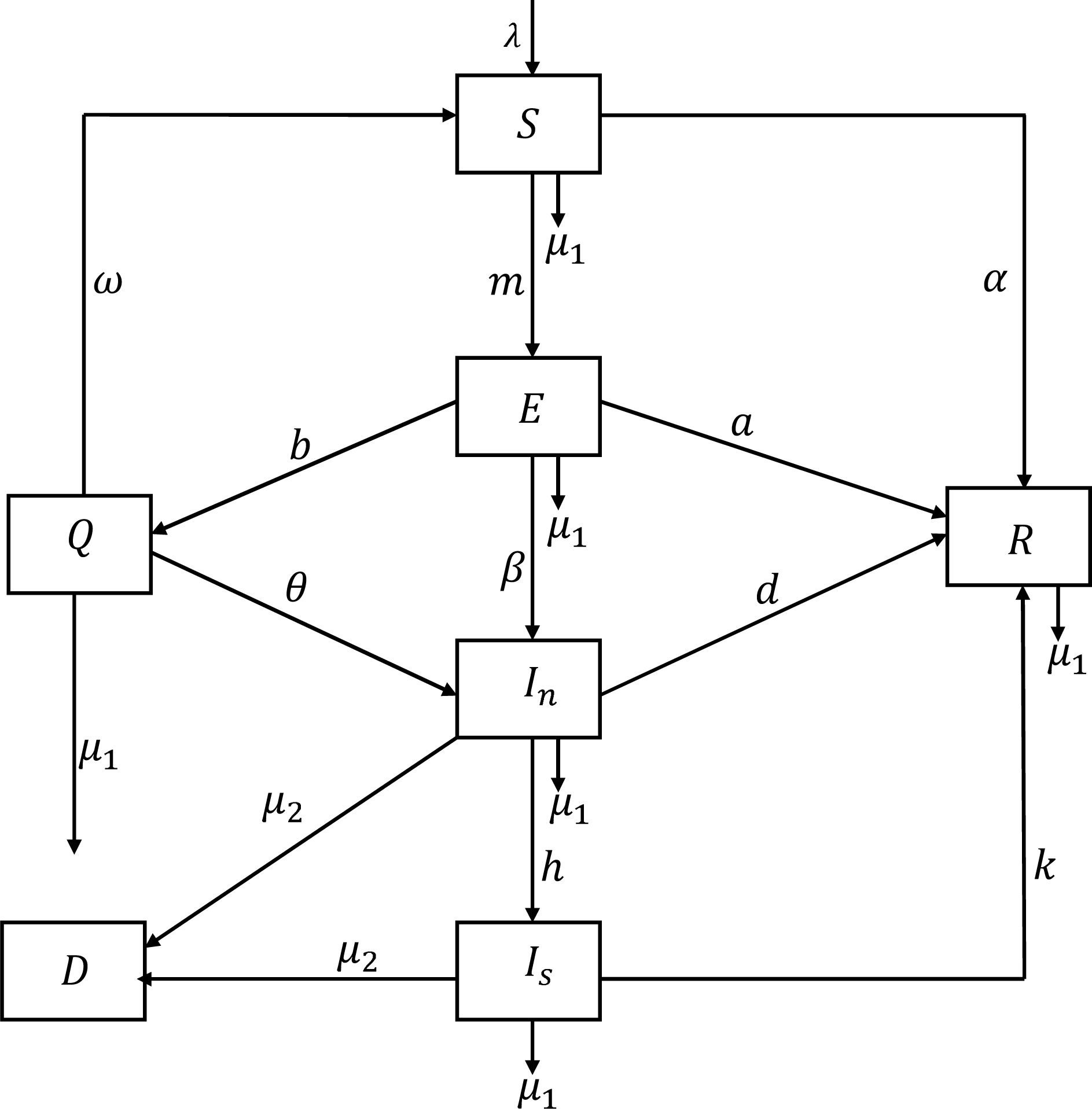
Schematic diagram for the model

The model is developed under the following mathematical assumptions

▪ The recovered individuals do not become susceptible to the disease again.
▪ Potion of the infected individuals and isolated individuals can die due to disease.
▪ Every susceptible individual has equal chance of being infected.
▪ New recruit enters the population through birth and migration.
▪ The quarantined individuals can be infected when they come in contact with contaminated surface or infected person.

Based on the mathematical assumptions and the schematic diagram, we formulate the following differential equations.

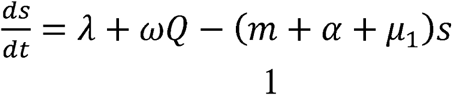

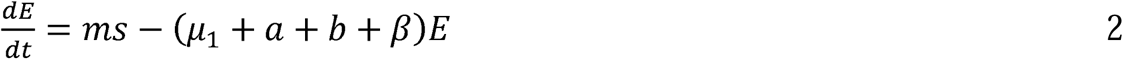

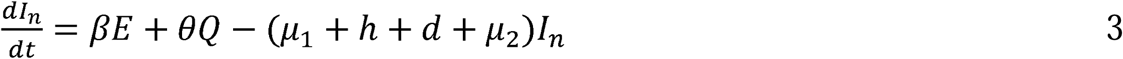

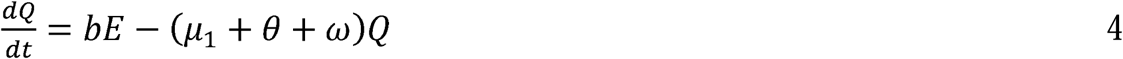

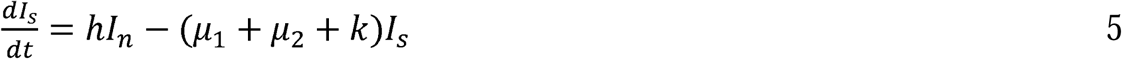

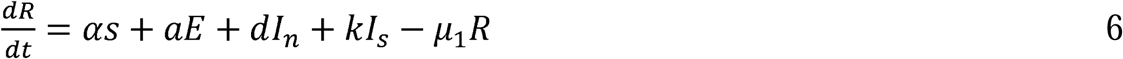

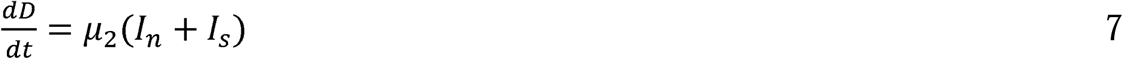

Where *N*(*t*) = *S*(*t*) + E(*t*) + *I*_*n*_(*t*) + *Q*(*t*) + *I*_*s*_(*t*) + *R*(*t*) + *D*(*t*)

## 3.0 Qualitative Analysis Of The Model

In this section we carry out the model analysis such that the equilibrium state, disease free equilibrium, and endemic equilibrium in order to determine stability of the model.

### 3.1 EQUILIBRIUM STATE

At the equilibrium point we have.

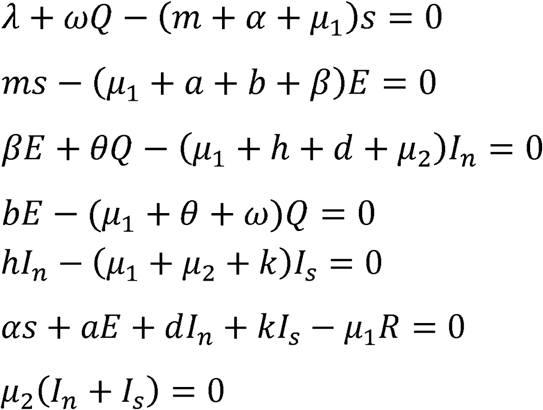

### 3.2 The Disease Free Equilibrium Of The Model

At the disease free equilibrium

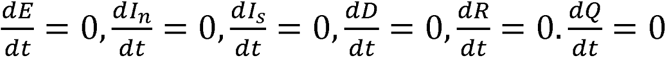

The disease free equilibrium of our model equation is

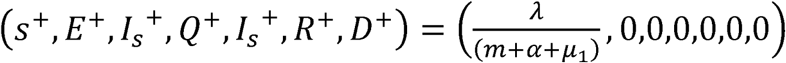

### 3.3 The Endemic Equilibrium Of State

The endemic state of our model equations is given below

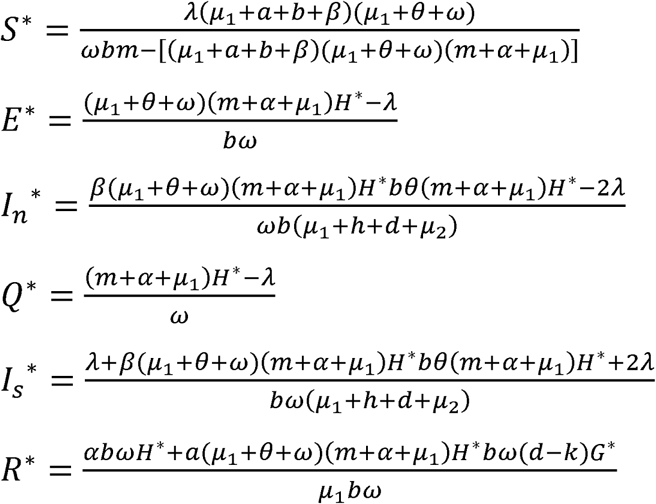

Where and *S*^*^ = *H*^*^ and *I*_*n*_^*^ = *G*^*^

### 3.4 The Basic Reproduction Number (*R*_0_)

In any disease model, the basic reproduction number (*R*_0_) is highly significant since it is the most relevant epidemiological distinguishing feature to determine the nature of disease. We therefore, use the next generation matrix approach to evaluate basic reproduction number of the model.

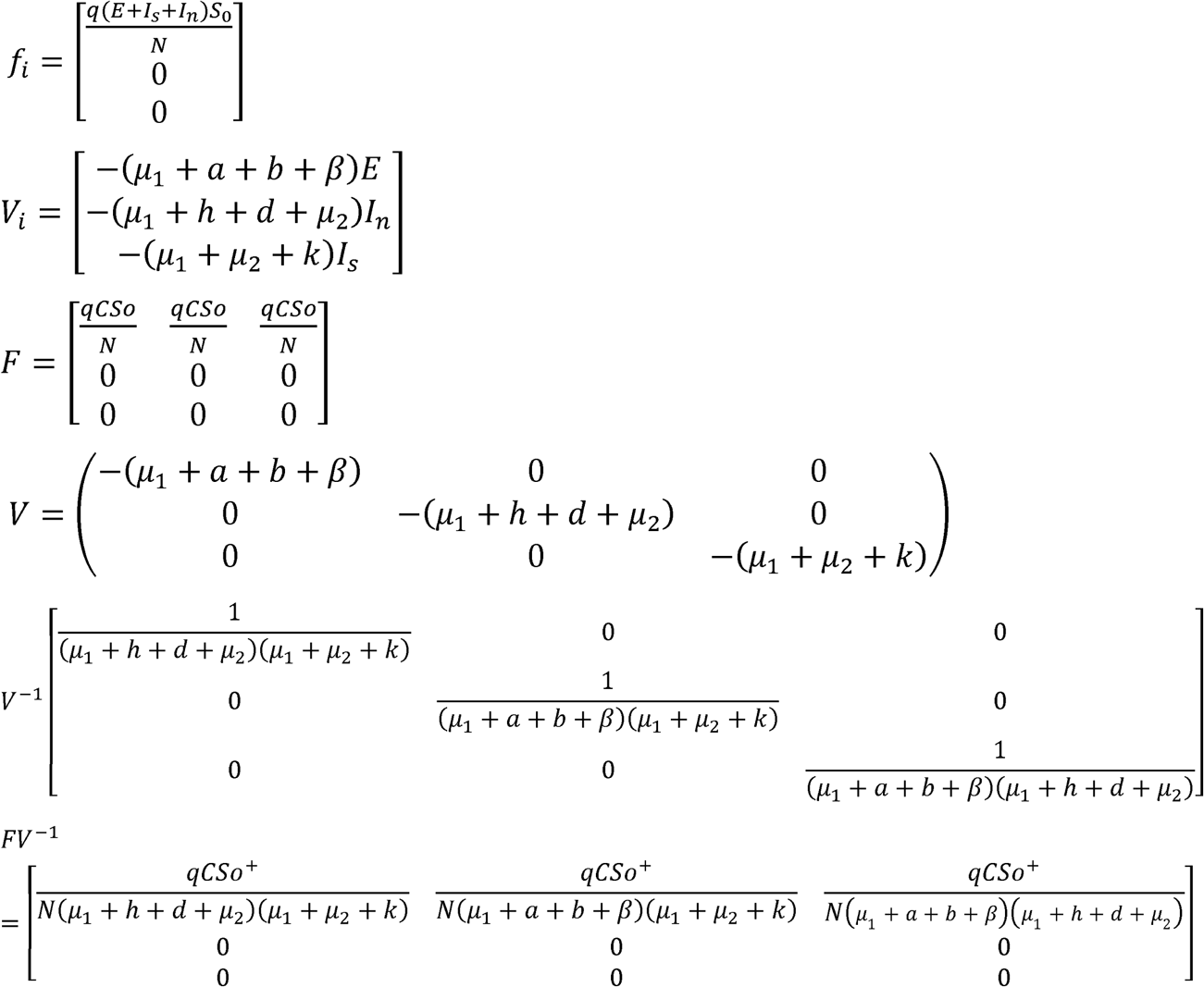

The basic reproduction number (*R*_0_) is the spectral radius of the matrix (*FV*^−1^) which is given by

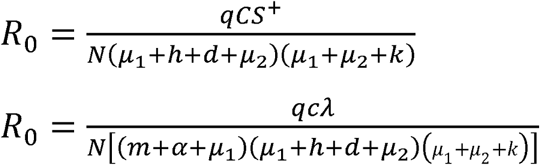

### 3.5 Local Stability Of Disease Free Equilibrium

Let *T* = *λ* + *ωQ* − (*m* + *α* + *μ*_1_)*S*

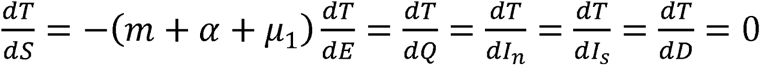

Let *A* = *ms* − (*μ*_1_ + *a* + *b* +*β*)*E*

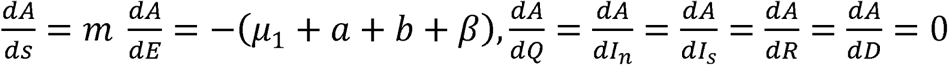

Let *B* = *βE* + *θQ* − (*μ*_1_ + *h* + *d* + *μ*_2_) *I*_*n*_

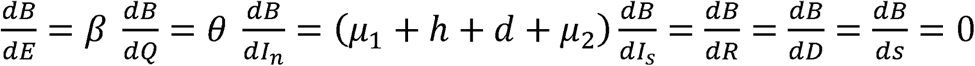

Let *F*= *bE* − (*μ*_1_ + *θ* + *ω*)*Q*

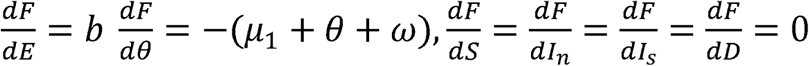

Let *V*= *hI*_*n*_ − (*μ*_1_ + *μ*_2_ + *k*)*I*_*s*_

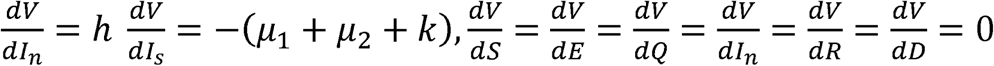

Let *X*= *αs + aE + dI*_*n*_ *+ KI*_*s*_ − *μ*_1_*R*

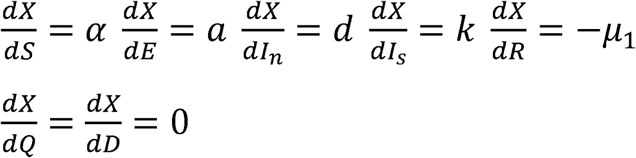

Let *Y*= *μ*_2_(*I*_*n*_ + *I*_*s*_)

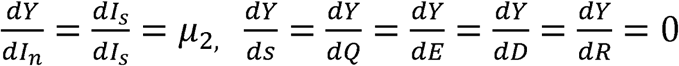

The Jacobian matrix for the model is given below

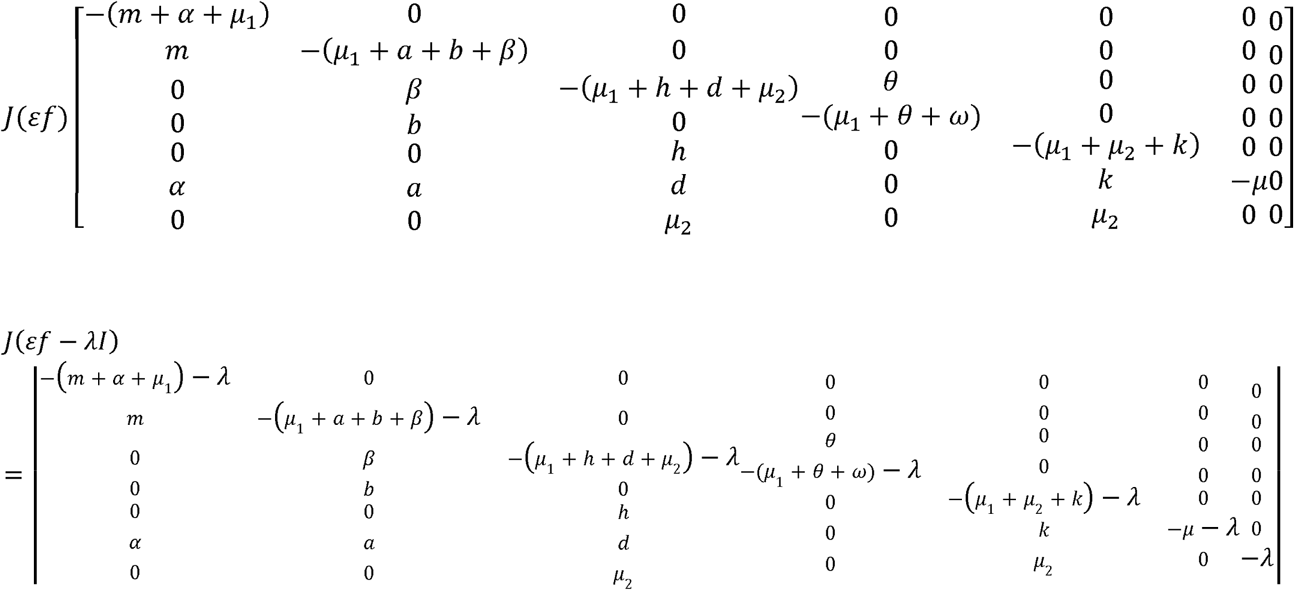

The characteristic equation is

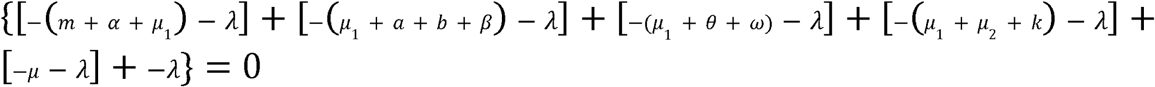

Since all the eigen values are negative, we conclude that the system is locally and asymptotically stable around its disease free equilibrium.

### 4.6 Numerical Simulation

We perform numerical simulation of the model in order to compare the results of our model with the real data obtained from published articles by WHO and other researchers, considering a total of 56,956 confirmed cases in Nigeria. Figures 1,2,3,4,5,6,7 and 8 are very useful to this work and their significance have been depicted under each of the figures. The numerical simulation of our model is performed using MATLAB and set of parameter values below:

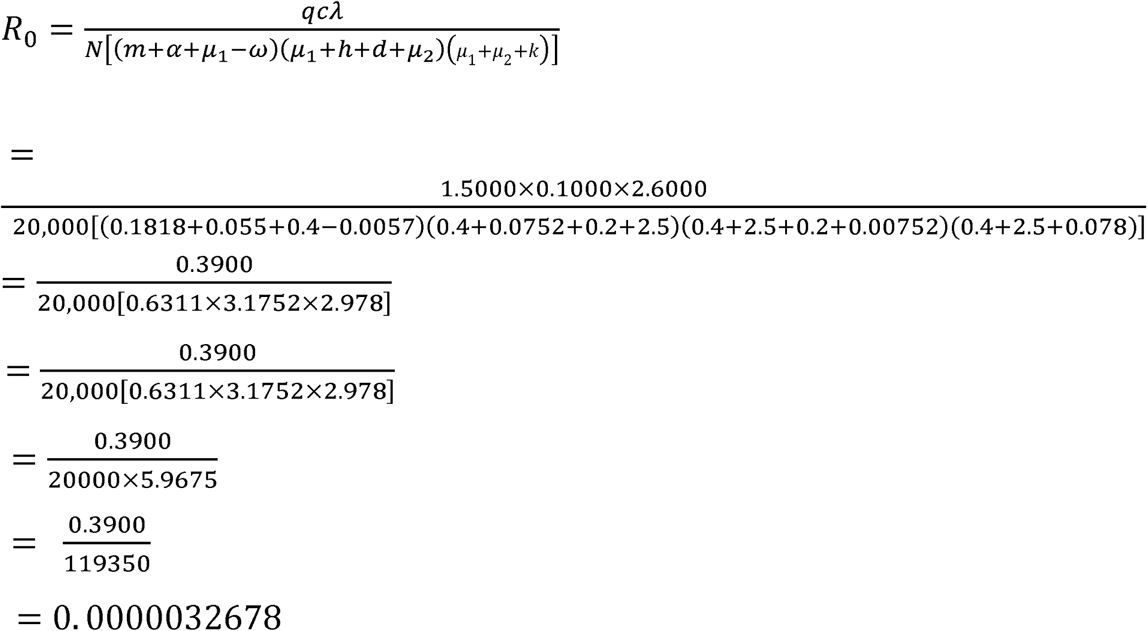

| Parameter | Values | Description | source |
| --- | --- | --- | --- |
| 1. $m$ | 0.1818 | Rate at which susceptible individuals become exposed | [15] |
| 2. $\beta$ | 0.1493 | Rate at which exposed individuals become infected | [13] |
| 3. $h$ | 0.0752 | Isolation rate | [15] |
| 4. $\mu_2$ | 2.5000 | Disease induced death rate | Estimated |
| 5. $\mu_1$ | 0.0400 | Natural mortality rate | Estimated |
| 6. $b$ | 2.5000 | Quarantine rate | Estimated |
| 7. $\theta$ | 0.1500 | Rate at which quarantined people become infected | Estimated |
| 8. $a$ | 0.0058 | Rate at which exposed people get recovered | [11] |
| 9. $\alpha$ | 0.0550 | Recovering rate of susceptible individuals | Estimated |
| 10. $\omega$ | 0.0057 | Rate at which quarantined people become susceptible | Estimated |
| 11. $k$ | 0.0780 | Rate at which Isolated people get recovered | Estimated |
| 12. $\lambda$ | 2.6000 | Recruitment rate | Estimated |
| 13. $d$ | 0.2000 | Recovering rate of infected individuals | Estimated |
| 14. $q$ | 1.5000 | Per contact rate of infection | Estimated |
| 15. $c$ | 0.1000 | Probability of getting COVID-19 | Estimated |

**Fig (2).**
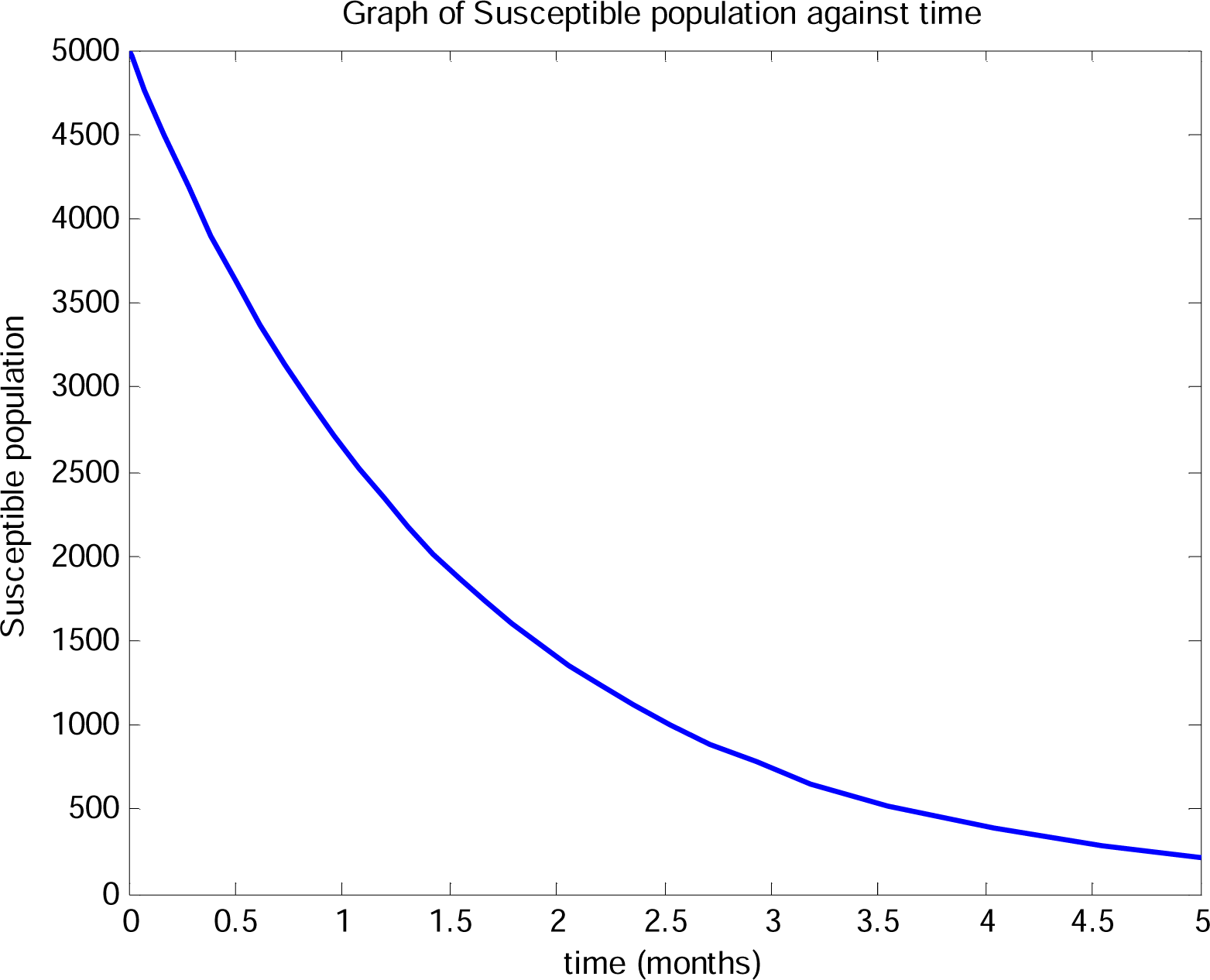
is the graph of susceptible population against time. We observed that the susceptible individuals decrease rapidly as time increases showing that few numbers will be susceptible to the disease and the infection would be eradicated from Nigeria as time progresses.

**Fig (3).**
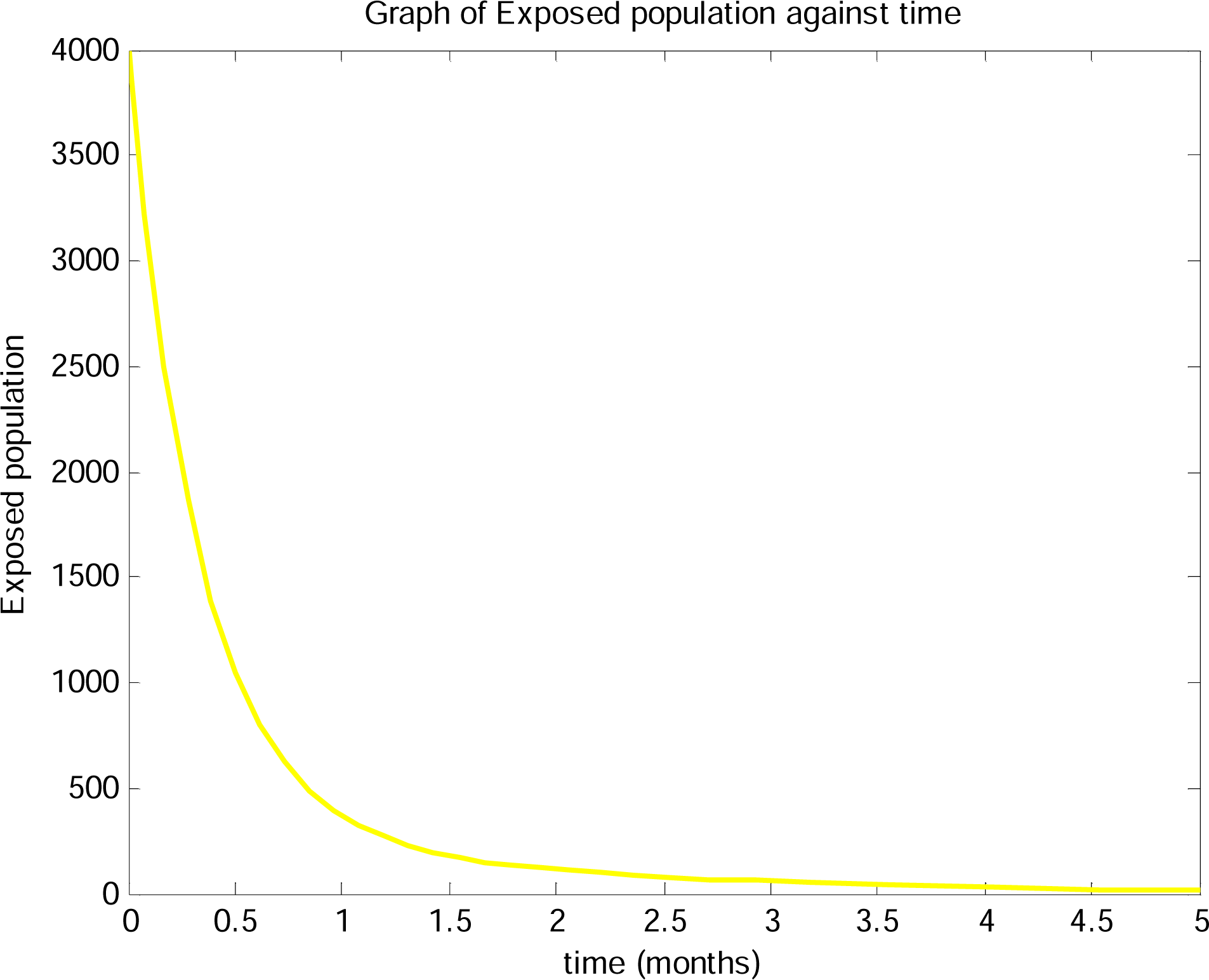
is the graph of exposed population against time, from the graph, the exposed population decreases as time increases, the exposed population decreases to zero indicating that with time the COVID-19 pandemic can be controlled in Nigeria since the exposed population tends to zero.

**Fig (4).**
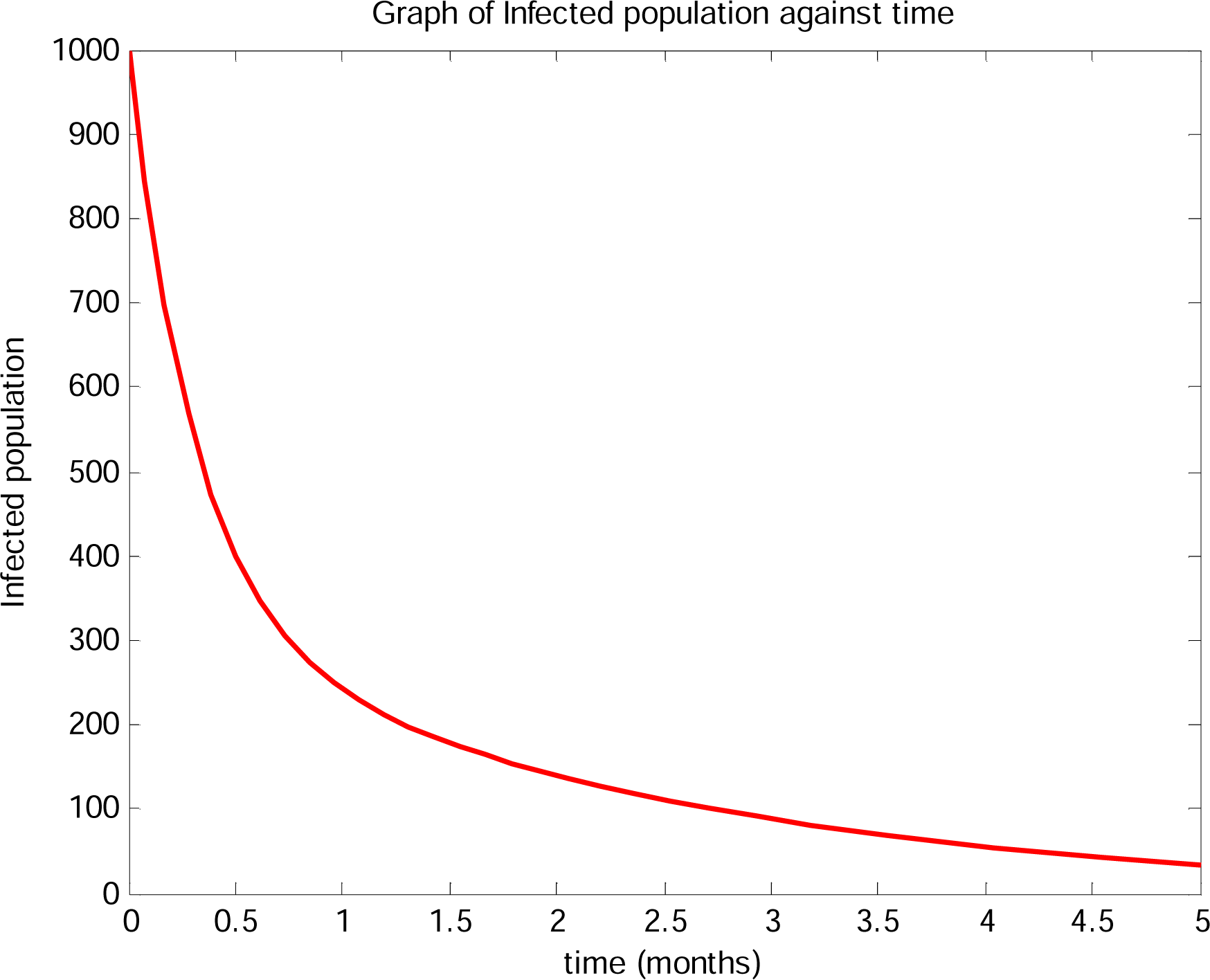
is the graph of infected population against time, it can be seen from the graph that infected population decreases monotonically as time increases which means that with time no one will be infected and the disease dies gradually from Nigeria.

**Fig(5).**
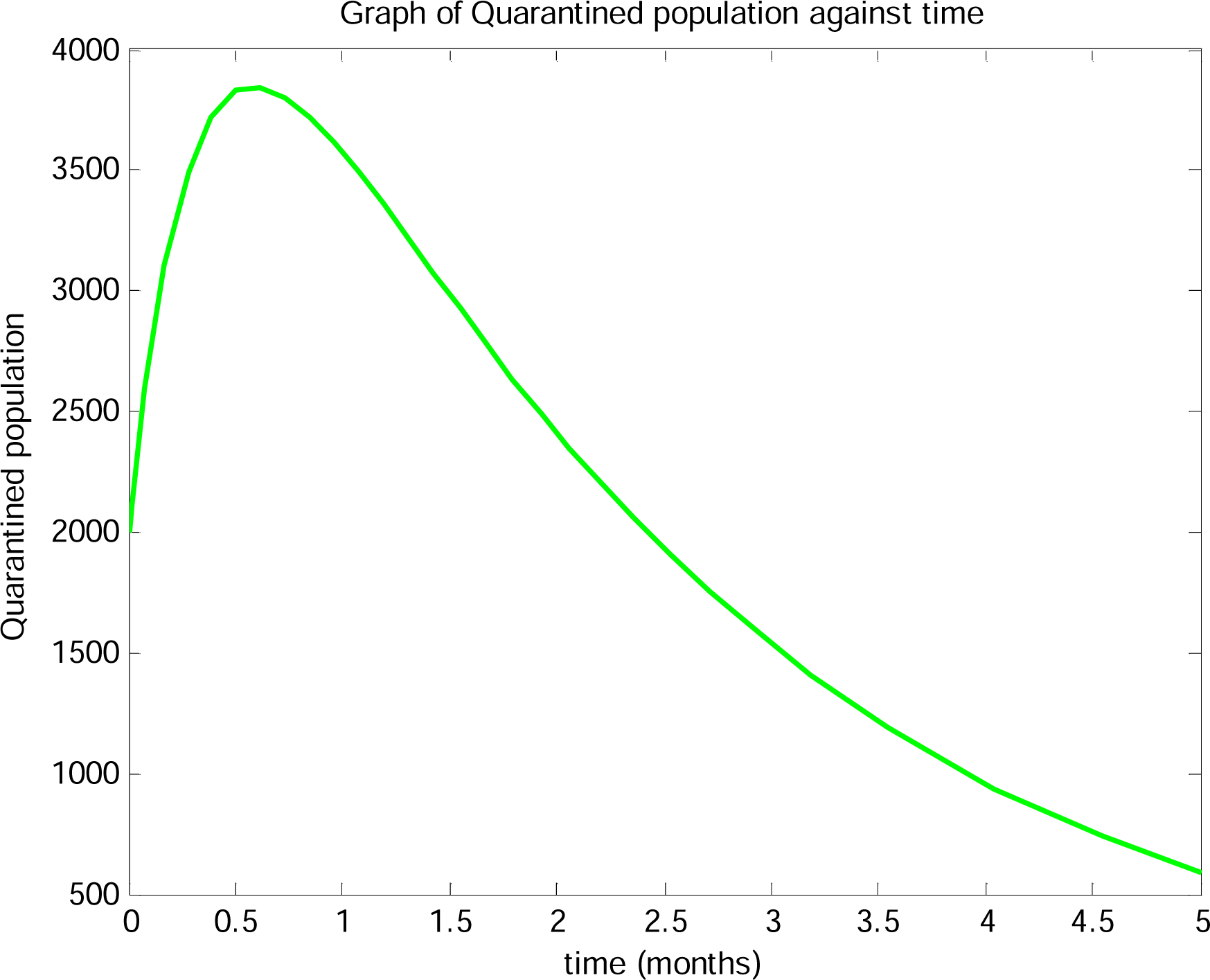
is the graph of quarantined population against time, initially quarantined population increases rapidly, the initial exponential growth could be as a result of government intervention measures such as media cover-age on social distancing and period of lockdown, it later decreases due to decrease in susceptible and infected population.

**Fig (6).**
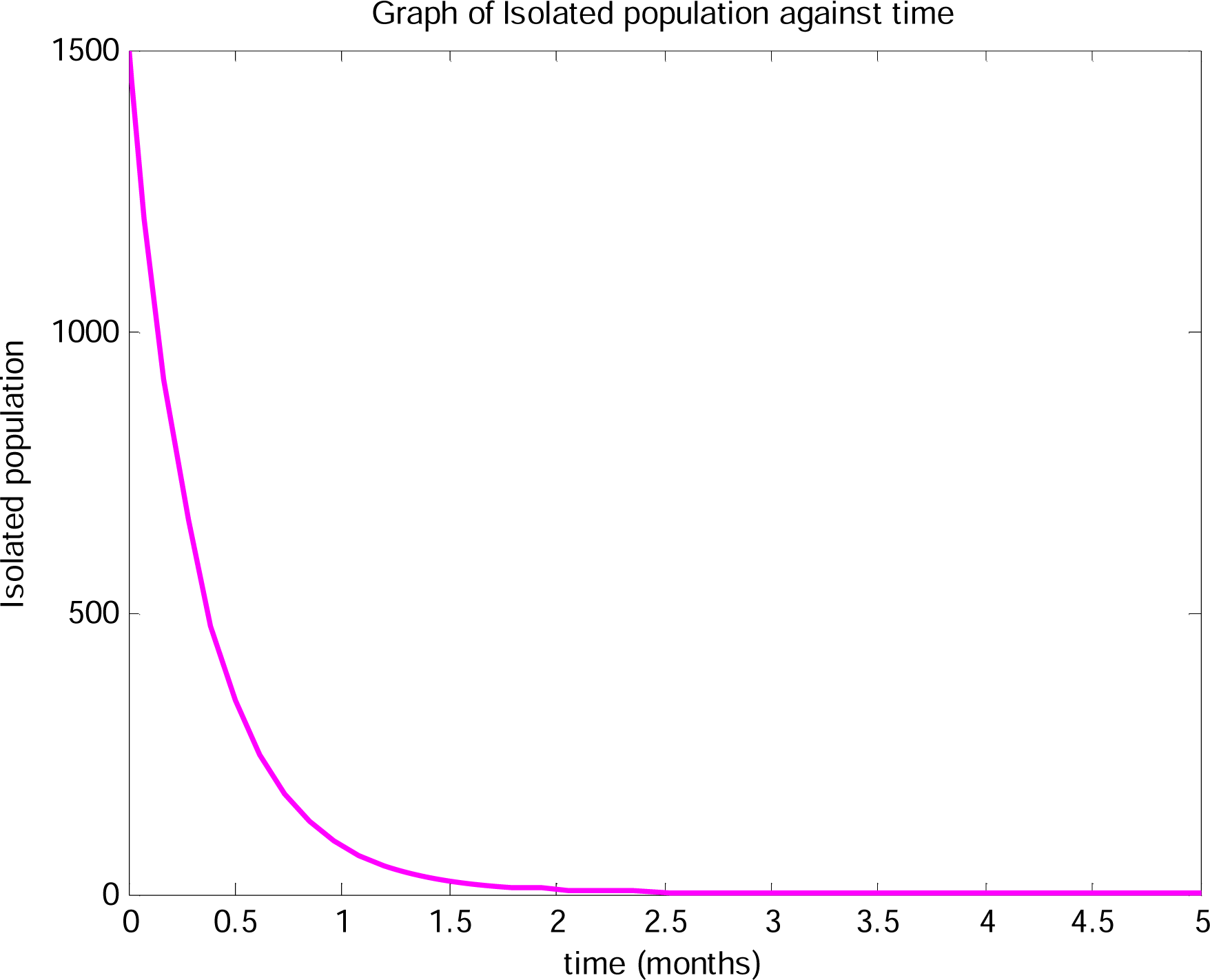
is the graph of isolated population against time, it is observed that isolated population decreases in Nigeria as time increases this could be as a result of decease in infected population and rapid increase of recovered individuals.

**Fig(7).**
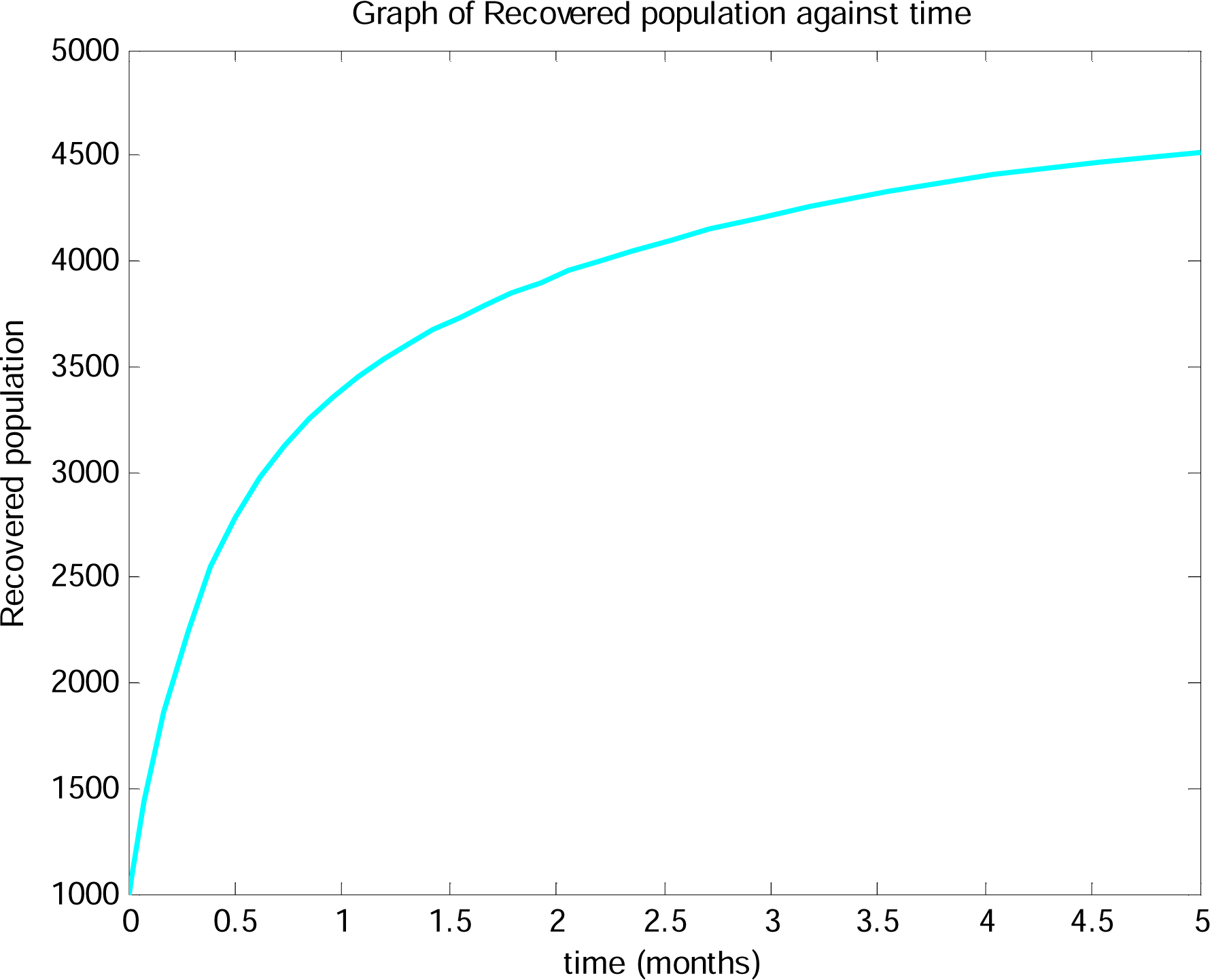
is the graph of recovered population against time, the recovered individuals incease greatly as time increases, this could be as a result of effective isolation and decrease in rate of infection.

**Fig (8).**
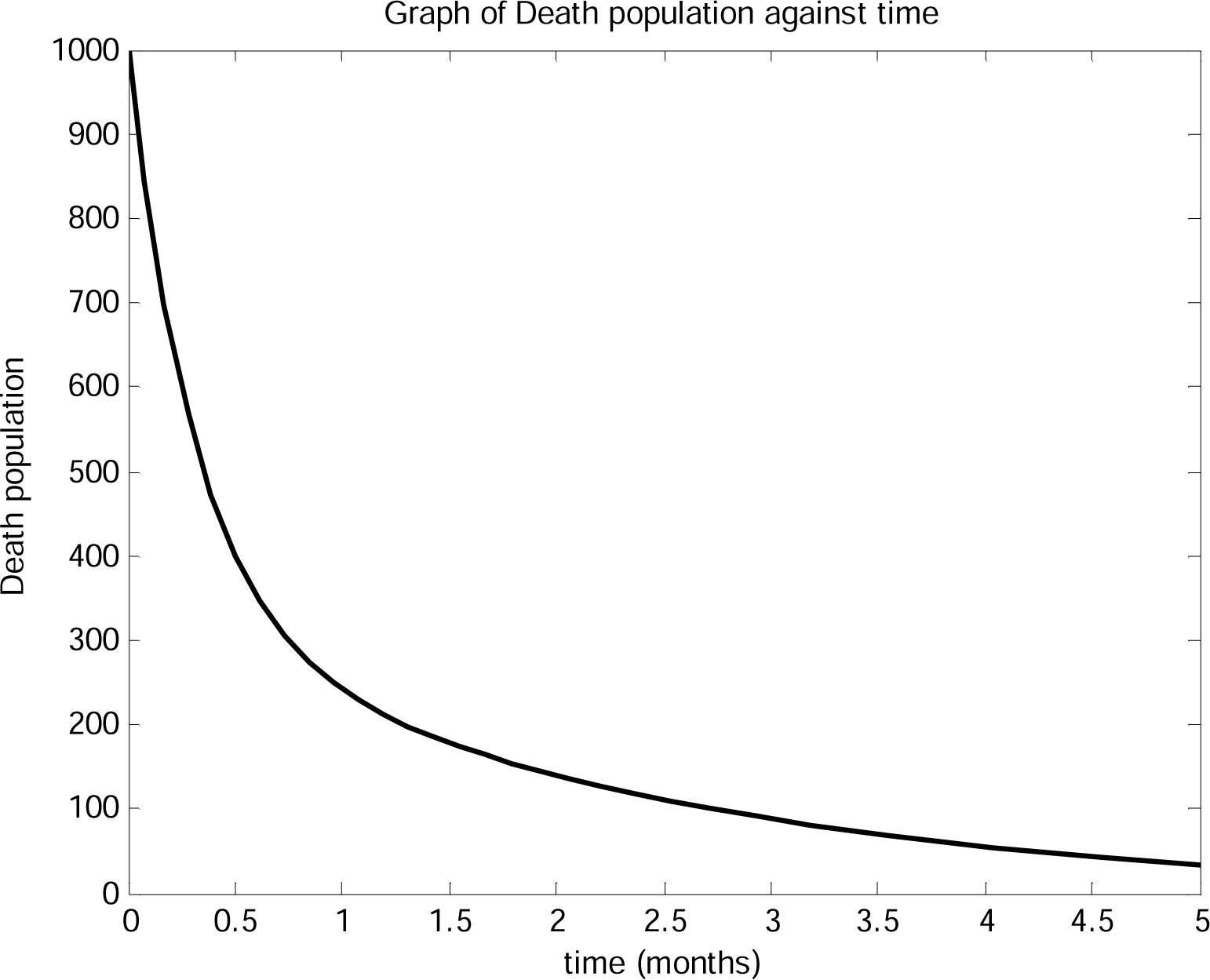
is graph of death population against time in Nigeria, the population is at high rate when the outbreak occurs initially but later decreases with time showing that disease induced death tends to zero as time progresses.

## 5.0 Conclusion

COVID-19 pandemic is the second world disaster after the World War II, government of Nigeria has taken some policies to control the catastrophic nature of the deadly disease in their own capacity. In this article, we have developed and analyzed a mathematical model which incorporates isolation and quarantine strategies as control measures. Analysis of the model shows that diseases free equilibrium is locally and asymptotic stable. The numerical simulation and analysis of the model revealed that COVID-19 pandemic can be eradicated from the population.

## Data Availability

The data used are gotten from some publications and some realistically estimated

